# Predicting the Risk of Asthma Development in Youth Using Machine Learning Models

**DOI:** 10.1101/2024.06.24.24309438

**Authors:** Matthew Xie, Chenliang Xu

## Abstract

Asthma is a chronic respiratory disease characterized by wheezing and difficulty breathing, which disproportionally affects 4.7 million children in the U.S. Currently, there is a lack of asthma predictive models for youth with good performance. This study aims to build machine learning models to better predict asthma development in youth using easily accessible national survey data. We analyzed cross-sectional combined 2021 and 2022 National Health Interview Survey (NHIS) data from 9,716 youth subjects with their corresponding parent information. We built several machine learning models with various sampling techniques (under- or over-sampling) for asthma prediction in youth, including XGBoost, Neural Networks, Random Forest, Support Vector Machine (SVM), and Logistic Regression. We examined the associations of potential risk factors identified from both Random Forest and Least Absolute Shrinkage and Selection Operator (LASSO) with asthma in youth. Between the different sampling techniques, undersampling the major class (subjects without asthma) yielded the best results in terms of the area under the curve (AUC) and F1 scores for the different predictive models. The Logistic Regression performed the best with the under-sampled data, yielding an AUC score of 0.7654 and an F1 score of 0.3452. In addition, we have identified additional important factors associated with asthma development in youth, such as low family poverty ratio and parents ever had asthma. This study successfully built machine learning models to predict asthma development in youth with good model performance. This will be important for early screening and detection of asthma in youth.

## Introduction

Asthma is a chronic lung disease portrayed by deadly asthma attacks that result in wheezing, breathlessness, and chest tightness (1). In 2013, around $5.92 billion was spent on pediatric asthma in the U.S. (2). Currently, the only protective measures to combat asthma and asthma attacks are medications and avoidance of triggers (3, 4). The American Lung Association identified several factors that may cause asthma, such as family history, occupational exposure, and smoking (5). Furthermore, studies have shown that 80% of asthma cases arise during the first six years of a person’s life (6). Due to the lack of a cure, it is crucial that asthma development in youth can be predicted and detected early so that prevention and early intervention may take place.

The recent development of artificial intelligence techniques (especially machine learning and deep learning), as well as the data availability of detailed patient information, empowers the prediction and risk assessment of various chronic diseases. For example, machine learning has been employed in predicting cardiovascular diseases (7–9). While many machine learning models have been successfully built for predicting asthma exacerbations among asthmatic patients (10–16), fewer studies have been conducted on predicting asthma risk using various datasets especially publicly available data, such as the 2019 Michigan BRFSS data and a small dataset of 202 children from Ibn Sina Hospital Center in Morocco (7, 8). However, the dataset was either too small, limited to a specific region, or relied heavily on clinical factors, which may limit model performance and implementation (8). This is evident by the relatively poor model performance in the 2019 Michigan BRFFS data, which employs two sampling techniques, Synthetic Minority Over-Sampling Technique (SMOTE) and Random Over-Sampling Examples (ROSE), resulting in the highest AUC score of 0.63 with the logistic regression. These predictive models’ performance could be improved by using a larger dataset and employing more advanced data processing and machine learning techniques.

This study aims to build predictive models for asthma development in youth using the combined National Health Interview Survey (NHIS) 2021 and 2022 survey data, which ensures a large sample size. Furthermore, we employed different sampling techniques, including oversampling, undersampling, and both, to address the class imbalance issue (most survey subjects had no asthma). Two complementary methods, Least Absolute Shrinkage and Selection Operator (LASSO) and random forest models, were used to identify factors associated with asthma development in youth. Finally, different machine learning models, such as logistic regression and neural networks, were used for predicting asthma development in youth, and their model performances were compared.

## Methods

### Data source

This study used the combined cross-sectional 2021 and 2022 NHIS children and adults data (9). The NHIS is a data collection program for the National Center for Health Statistics (NCHS), a part of the Centers for Disease Control and Prevention (CDC). The NHIS is a household interview survey that involves face-to-face interviews followed by telephone interviews throughout the year. The NHIS collects data about the health of the noninstitutionalized civilian population of all ages in the United States. The purpose of the NHIS is to monitor the health of the United States population by categorizing health trends by demographic and socioeconomic circumstances. The NHIS 2021 and 2022 children and adults data are publicly available from the CDC website (9).

### Data pre-processing

The outcome variable of this study is asthma, which is based on the question, “Has a doctor or other health professional EVER told you that you had asthma?” If the answer is “Yes,” the subject is considered to have asthma. If the answer is “No,” the subject is regarded as having no asthma.

As shown in Appendix Figure 1, we merged the youth survey data with their linked parent data by matching the unique child’s household ID with their respective parents, which was conducted for both 2021 and 2022 NHIS data, respectively. The merged 2021 NHIS dataset has 7,070 youth subjects, while the 2022 NHIS dataset has 6,261 youth subjects with their corresponding parent information. We combined the 2021 and 2022 NHIS data to increase the sample size to form the final dataset with 13,331 youth subjects.

To get the data ready for the machine learning models, we employed several preprocessing or cleaning steps (Appendix Figure 2). According to the codebook for adult and youth NHIS data, we first selected relevant variables potentially related to asthma development according to previous literature (10–14) and excluded irrelevant variables in the subsequent analysis. For example, questions specific to the survey design or a specific population not related to asthma, such as cancer patients, were not included in the analysis. After the irrelevant variables were removed, we removed all variables with only one level (constant variables) or variables only focused on a subgroup of age (such as age 5-17) because this study focuses on youth aged 0-17. To avoid potential multicollinearity problems, we examined the multicollinearity issues using the variance inflation factor (VIF) values. We removed highly correlated variables with VIF values larger than 2. Once this step was completed, we deleted all records with missing values for the remaining variables and responses such as “don’t know,” “refused,” or “not ascertained.” The final dataset contains 9,716 youth subjects with their corresponding parent information, which we used for subsequent analysis.

### Feature/variable selection

To identify important features for predicting asthma in youth, we employed two complementary machine learning models (LASSO and Random Forest) for feature selection (Appendix Figure 2). LASSO selects important features by first establishing a penalized regression model where the dependent variable is equal to the sum of the independent variables multiplied by the estimated coefficients and the error term. LASSO essentially finds the coefficient values (while shrinking them toward zero) that minimize the sum of the squared differences between predicted and actual values (15). This process makes LASSO useful for removing irrelevant features (16). Random forest can be used for both classification and regression (17). The random forest model evaluates the variables based on the aggregated outputs from individual decision tree models to select the most relevant features (17). Each model generated a list of the most significant variables or features for asthma prediction. We used the mean of feature importance values for all variables as the cutoff to select the variables/features. There were 34 variables selected from LASSO, 13 of which were also chosen by Random Forest. To maximize the model performance, we utilized the union (34 variables) of two variable lists for machine learning models. However, to understand how these variables contribute to asthma prediction in youth, we took the intersection (13 variables) of the two lists to minimize the false positive.

### Class imbalance

Considering that the proportion of youth subjects with asthma in the dataset is relatively small, the class label for asthma in our dataset is unbalanced, with most subjects having no asthma, which can significantly influence the model performance of predictive models. To deal with the class imbalance, after splitting the data into training and test datasets with a ratio of 7 to 3, we applied different sampling techniques to balance the class labels in the training dataset. The sampling techniques include SMOTE-Tomek (both oversampling and undersampling), undersampling, and oversampling (SMOTE) (18). The undersampling technique will randomly select some samples from the major class (subjects without asthma) so that the number of subjects with asthma is similar to those without asthma. As an oversampling technique, SMOTE will create additional synthetic records for subjects with asthma so that the numbers of subjects with and without asthma are similar. SMOTE-Tomek is a sampling technique employing both oversampling and undersampling (19). This combination of over and undersampling helps balance the cons of only performing under or oversampling.

### Building machine-learning models

In this study, we used multiple machine-learning models such as random forest, XGBoost, neural networks, support vector machine with a linear kernel, and logistic regression to predict asthma development in youth (Appendix Figure 3).

### Random Forest

Random forest is a machine-learning modeling technique that can be used in both classification and regression (17). A typical random forest model is composed of numerous decision trees, each of which classifies data points into different class labels depending on their features. Eventually, the random forest model takes the output of each decision tree model and chooses the best output. This model is good for large datasets such as the NHIS dataset and has high precision (20).

### XGBoost

Similar to the tree-based random forest model, Extreme Gradient Boosting is a gradient-boosted decision tree (21). However, instead of outputting the average of each decision tree output in the random forest, XGBoost outputs the weighted average of each decision tree output. XGBoost is characterized by a combination of weak decision tree models to create a strong combined model. An advantage of XGBoost is that it reduces bias and underfitting (21).

#### Feedforward Neural Networks

A perceptron is a machine learning algorithm that has many nodes whose output is an input for another node (22). Within each node, the neural network takes the sum of the product of the weight and value for each input, adds a bias value, and passes it over to an activation function. The resultant value is then passed to nodes in the next layer in the network. The high computational power and accuracy of feedforward neural networks make it desirable for this study (23).

#### Support Vector Machine

Support vector machine (SVM) is mainly used for classification but can also be used for regression analysis (24). SVM involves the creation of a hyperplane or line that the model uses to distinguish between different groups of data points. After the SVM is run, the model outputs a number either greater than 1 or less than –1, with each scenario classifying the data point as a group. SVM is typically used for small datasets due to its high accuracy and long run times (25). In this study, the linear kernel was used.

#### Logistic Regression

A logistic regression model uses the sigmoid function to convert the data to a probability and assigns the data to groups based on the probabilities (26). The logistic regression typically outputs the likelihood of a data point belonging to a particular group, making it good for predictions in the case of asthma.

The predictive models were trained on the training dataset with no sampling, oversampling, undersampling, and combination of oversampling and undersampling. We measured the model performance on the test dataset using the F1 score, area under the curve (AUC), precision, accuracy, and sensitivity.

### Statistical analysis

To measure the association of independent variables with asthma development in youth, we selected the variables in the intersection of the two variable lists identified from LASSO and Random Forest. We conducted two types of statistical analysis, namely a Chi-square test for categorical variables and a two-sample *t-*test for numerical variables, to determine if the variables were significantly associated with asthma development in youth.

## Results

### Feature selection

After data pre-processing and initial variable selection, the 2021 and 2022 combined NHIS datasets contained 9,716 youth subjects with 39 variables. Among 9,716 youth subjects, 1,043 subjects (10.7%) have reported asthma, and 8,673 subjects (89.3%) do not have reported asthma. Two different but complementary feature selection models were employed to identify important variables that might be associated with asthma development in youth: LASSO and Random Forest. As shown in Appendix Table 1, using LASSO, 34 variables were identified to be important for asthma prediction, such as age, gender, and whether adults ever had asthma. Through Random Forest, 13 variables were identified, such as age, gender, and family poverty ratio, which are also included in the variable list from LASSO (Appendix Table 1). To maximize the predictive ability of machine learning models and minimize the false negatives, we took the union of selected variables from two lists, which resulted in 34 variables in total.

**Table 1.** Model performance measures of machine learning models for asthma with different sampling techniques.

| Sampling Technique | Predictive Model | AUC score | Precision | Sensitivity | Accuracy | F1 Score |
| --- | --- | --- | --- | --- | --- | --- |
| No sampling | Random Forest | 0.7460 | 0.4706 | 0.0252 | 0.8906 | 0.0478 |
|  | XGBoost | 0.6796 | 0.3358 | 0.1447 | 0.8755 | 0.2022 |
|  | Neural Network | 0.5449 | 0.3711 | 0.1132 | 0.8823 | 0.1735 |
|  | Logistic Regression | 0.7689 | 0.4082 | 0.0629 | 0.8878 | 0.1090 |
|  | SVM (linear) | 0.7503 | 0.0000 | 0.0000 | 0.8909 | 0.0000 |
| Undersampling | Random Forest | 0.7461 | 0.2157 | 0.6384 | 0.7074 | 0.3225 |
|  | XGBoost | 0.7067 | 0.1922 | 0.6384 | 0.6679 | 0.2955 |
|  | Neural Network | 0.6640 | 0.2602 | 0.5031 | 0.7897 | 0.3430 |
|  | Logistic Regression | 0.7654 | 0.2325 | 0.6698 | 0.7228 | 0.3452 |
|  | SVM (linear) | 0.7589 | 0.2273 | 0.6541 | 0.7197 | 0.3374 |
| Oversampling | Random Forest | 0.7483 | 0.4032 | 0.0786 | 0.8868 | 0.1316 |
|  | XGBoost | 0.6838 | 0.3893 | 0.1604 | 0.8810 | 0.2272 |
|  | Neural Network | 0.5874 | 0.2459 | 0.2799 | 0.8278 | 0.2618 |
|  | Logistic Regression | 0.6316 | 0.2315 | 0.4434 | 0.7787 | 0.3042 |
|  | SVM (linear) | 0.6997 | 0.2398 | 0.4245 | 0.7904 | 0.3065 |
| SMOTE-TOMEK | Random Forest | 0.7492 | 0.4333 | 0.0818 | 0.8882 | 0.1376 |
|  | XGBoost | 0.6738 | 0.3261 | 0.1415 | 0.8744 | 0.1974 |
|  | Neural Network | 0.5835 | 0.2890 | 0.2390 | 0.8528 | 0.2616 |
|  | Logistic Regression | 0.6949 | 0.2331 | 0.4434 | 0.7801 | 0.3055 |
|  | SVM (linear) | 0.6996 | 0.2394 | 0.4245 | 0.7901 | 0.3061 |

### Predictive models for asthma development in youth

Table 1 summarizes the performances of the five predictive models, including random forest, XGBoost, neural networks, logistic regression, and support vector machine (SVM) with the linear kernel. Trained with the original unbalanced data without any sampling techniques, XGBoost showed the best model performance with AUC and F1 scores of 0.6796 and 0.2022. In contrast, while it has the highest accuracy, SVM (linear) had no precision and sensitivity, especially the F1 score, indicative of poor model performance. The low F1 score for the models may be due to the class imbalance issue, leading to predicting every subject as not having asthma.

We applied the undersampling technique to address the class imbalance issue (only 10.3% of subjects with asthma) in the dataset, which randomly selected the same number of samples from the major class (subjects without asthma) as the minor class (subjects with asthma). As shown in Table 1, all machine learning models trained on the balanced dataset with undersampling performed much better than those trained on the original data, as evidenced by the model performance measures, specifically the F1 score (the most appropriate model performance measure for class imbalance). For example, while the AUC and F1 scores for the neural networks model trained on original data are 0.5449 and 0.1735, the model performance was significantly increased when the model was trained on the under-sampled dataset, with the AUC and F1 score of 0.6640 and 0.3430. In addition, trained on the balanced dataset, the logistic regression (AUC = 0.7654 and F1 = 0.3452) and support vector machine (AUC = 0.7589 and F1 = 0.3374) performed well based on their AUC and F1 scores.

To deal with the class imbalance, another sampling technique, oversampling, was applied to oversample the minority group (subjects with asthma). Based on the F1 score and AUC value, as shown in Table 1, the oversampling technique improved the model performance compared to the models trained on the original dataset. For example, the AUC and F1 scores for the neural networks model are 0.5874 and 0.2618. However, the performance of the models trained on the oversampled dataset was poorer than that of those trained on the under-sampled dataset. By comparison, the support vector machine model performed the best among the five models trained on the oversampling dataset, with an F1 score of 0.3065 and an AUC of 0.6997.

The combination of both oversampling and undersampling using SMOTE-Tomek technique was applied to the training dataset to balance potential issues with either oversampling alone or undersampling alone. As shown in Table 1, while the model performance for all models is better than the models trained on the original dataset, the F1 scores are similar to models trained on the oversampling dataset but poorer than models trained on the undersampling dataset. For example, the F1 score for the neural networks model trained on SMOTE-Tomek dataset is 0.2616.

Out of all the sampling techniques and models, the models trained on the undersampling of the major class yielded the best model performance. In addition, logistic regression and support vector machine were the two models with the best model performance based on the AUC and F1 scores.

### Potential risk factors for asthma development in youth

While it is important to predict the risk of asthma development in youth using machine learning models, it is equally important to identify factors that might contribute to asthma development, which can be valuable for early prevention. Using Random Forest and LASSO, we identified two lists of variables that might be the potential risk factors for asthma development. To test how these candidate variables are associated with asthma development, we selected the intersection of variables between two lists to avoid possible false positives and tested for their effects on asthma development. As shown in Table 2, as indicated by the significant P-value (P < .001), the occurrence of asthma is highly correlated with sex, age, the child’s health status, number of times the child visited urgent care in the past twelve months, number of times the child visited the emergency room in the past twelve months, if the child took prescription medication in the past twelve months, received a flu vaccine in the past twelve months, the asthma status of the child’s parents, the poverty level of the child’s family, and the symptoms of COVID-19. Based on the data shown in Table 2, males are more likely to develop asthma, children around age 11 are more likely to develop asthma than children around age 9, children with fair to poor health are more prone to asthma, children who visited the emergency room or urgent care more than 3 times are likely to have asthma, children whose parents had asthma are more likely to have asthma.

**Table 2.** Summary statistics on the variables identified by both LASSO and Random Forest.

| Variable | Asthma | No Asthma | Chi-square | P-value |
| --- | --- | --- | --- | --- |
|  | (n = 1043) | (n = 8673) |  |  |
| Sex |  |  | 33 | <.001 |
| Male | 616 (12.5%) | 4301 (87.5%) |  |  |
| Female | 427 (8.9%) | 4372 (91.1%) |  |  |
| Age (mean ± SD) | 11.3 ± 4.1 | 9.5 ± 4.7 | | <.001 |
| General Health Status |  |  | 420.9 | <.001 |

| Variable | Asthma | No Asthma | Chi- | P- |
| --- | --- | --- | --- | --- |
| Excellent | 416 (6.6%) | 5880 (93.4%) |  |  |
| Very Good | 327 (14.8%) | 1882 (85.2%) |  |  |
| Good | 233 (23%) | 780 (77%) |  |  |
| Fair | 60 (34.9%) | 112 (65.1%) |  |  |
| Poor | 7 (26.9%) | 19 (73.1%) |  |  |
| **Number of times visited urgent care in past 12 months** |  |  | 36.2 | <.001 |
| 0 times | 711 (9.8%) | 6538 (90.2%) |  |  |
| 1 time | 172 (12.3%) | 1230 (87.7%) |  |  |
| 2 times | 91 (13.7%) | 575 (86.3%) |  |  |
| 3 times | 30 (15.2%) | 167 (84.8%) |  |  |
| 4 times | 21 (17.8%) | 97 (82.2%) |  |  |
| 5+ times | 18 (21.4%) | 66 (78.6%) |  |  |
| **Number of times visited emergency room in past 12 months** |  |  | 79.9 | <.001 |
| 0 times | 824 (9.8%) | 7612 (90.2%) |  |  |
| 1 time | 144 (15.8%) | 770 (84.2%) |  |  |
| 2 times | 48 (17.4%) | 228 (82.6%) |  |  |
| 3 times | 14 (30.4%) | 32 (69.6%) |  |  |
| 4+ times | 13 (29.5%) | 31 (70.5%) |  |  |
| **Took prescription medicine in the past 12 months** |  |  | 440.2 | <.001 |
| Yes | 635 (20.3%) | 2490 (79.7%) |  |  |
| No | 408 (6.2%) | 6183 (93.8%) |  |  |
| **Received flu vaccine in past 12 months** |  |  | 7.05 | .008 |
| Yes | 538 (11.6%) | 4092 (88.4%) |  |  |
| No | 505 (9.9%) | 4581 (90.1%) |  |  |
| **Parent ever had asthma** |  |  | 142.2 | <.001 |
| Yes | 272 (20.1%) | 1083 (79.9%) |  |  |
| No | 771 (9.2%) | 7590 (90.8%) |  |  |
| **Family poverty ratio (mean ± SD)** | 3.5 ± 2.7 | 3.9 ± 2.9 | | <.001 |
| **Symptoms of COVID-19** |  |  | 31.5 | <.001 |
| No COVID-19 | 777 (10.2%) | 6877 (89.8%) |  |  |
| No Symptoms with COVID-19 | 50 (12.8%) | 342 (87.2%) |  |  |
| Mild Symptoms with COVID-19 | 134 (11.2%) | 1061 (88.8%) |  |  |
| Moderate Symptoms with COVID-19 | 63 (15.8%) | 336 (84.2%) |  |  |
| Severe Symptoms with COVID-19 | 19 (25.0%) | 57 (75.0%) |  |  |
| **Parent ever smoked a cigar** |  |  | 0.02 | 0.88 |
| Yes | 293 (10.6 %) | 2461 (89.4%) |  |  |
| No | 750 (10.8 %) | 6212 (89.2%) |  |  |
| **Urban-rural classification** |  |  | 1.67 | 0.64 |
| Large central metro | 299 (10.2%) | 2646 (89.8%) |  |  |
| Large fringe metro | 277 (11.1%) | 2216 (88.9%) |  |  |
| Medium and small metro | 320 (10.8%) | 2640 (89.2%) |  |  |
| nonmetropolitan | 147 (11.2%) | 1171 (88.8%) |  |  |
| **Household region** |  |  | 2.48 | 0.48 |
| Northeast | 151 (10.3%) | 1315 (89.7%) |  |  |
| Midwest | 204 (10%) | 1832 (90%) |  |  |
| South | 408 (11.3%) | 3211 (88.7%) |  |  |
| West | 280 (10.8%) | 2315 (89.2%) |  |  |

## Discussion

This study utilizes the data from the 2021 and 2022 youth and parent National Health Interview Survey to build machine-learning models to predict asthma development in youth. The NHIS includes questions ranging from pediatric to demographic and socioeconomic information. We linked the youth survey data with their parent survey data to account for the contribution of parents’ information to asthma development in youth. We applied several sampling techniques to deal with the class imbalance issue (most subjects without asthma), including oversampling, undersampling, and both. We built five different machine learning models to predict asthma development in youth. By comparison, predictive models trained on the dataset with the undersampling of the majority class yielded the best model performance overall. Undersampling outperformed no sampling and oversampling by balancing the training dataset without causing overfitting or creating synthetic data points that may not represent real-world scenarios. Among the models tested, the logistic regression and neural networks models were the best-performing models for predicting asthma development in youth, based on the AUC score and especially the F1 score. In addition, we found that several factors are significantly associated with asthma development in youth, such as age, gender, and family poverty ratio.

In this study, by employing different sampling techniques to address the class imbalance issue, we have successfully built multiple machine learning models to predict asthma development in youth aged 0 to 17 with good model performance. One previous study used machine learning to predict asthma amongst children aged 7 months to 12 years at a Morocco hospital, which achieved high performances with F1 scores around 0.80 (8). However, prenatal, perinatal, postnatal, and environmental factors cannot directly apply to the US population and children aged 13 to 17, which might limit its generalization. A recent study used Canadian Healthy Infant Longitudinal Development birth cohort data to predict pediatric asthma (27). The dataset was focused on children up to 4 years of age and included family medical history, clinical data, and environmental factors for young children. They used machine-learning models with 1,484 children and 132 variables and achieved great model performance with an AUC of 0.99 when predicting asthma in children at age 4 (27). However, their model cannot apply to children aged 5 to 17. Further, their data heavily relied on pediatric and prenatal clinical data extracted from patient health records and some environmental factors, which are difficult to obtain in real life, especially for those with low socioeconomic status. The dataset used in our study is national survey data containing basic demographics on youth aged 0 to 17, which is easily accessible, making it more practical.

In another asthma study conducted on the 2019 Michigan Behavioral Risk Factor Surveillance System (BRFSS) data, the group employed similar pre-processing techniques and used SMOTE and ROSE to deal with the class imbalance in the data (7). That study also employed similar machine-learning models on their dataset, yielding decent performance and identifying many risk factors. However, their dataset focuses on adults in Michigan rather than youth in the United States. Furthermore, the performance of the models in the Michigan study had the highest AUC score of 0.629 and F1 score of 0.287. In our study, the best model has an AUC score of 0.7632 and an F1 score of 0.3416. Therefore, the predictive models in our study performed much better in predicting asthma development in youth, which might be due to the improved pre-processing techniques and a more encompassing data source in this study. Some of the significant variables identified in the Michigan study were also identified in our study, such as flu vaccine and income. While the Michigan study showed that females have a higher risk for asthma development in adults, our study showed that males have a higher risk for asthma development in youth.

Besides building predictive models for asthma development in youth, another aim of this study is to identify potential risk factors for asthma development in youth, which might help with early asthma detection and prevention. Our study has identified several well-known risk factors for asthma development in youth, such as the presence of asthma in the parent, gender, socioeconomic status, and maternal smoking (28–31). It is well-known that family history is one of the major causes of asthma (5, 28). This could be due to the shared environment between the parents and child that may influence the development of asthma (32). Our study also found that boys are at a higher risk for asthma development than girls, which is also consistent with previous studies (29, 30). The increased risk for asthma development in boys might be due to the increased allergic inflammation and serum IgE levels in boys, as well as the smaller airway diameter relative to lung volumes in boys than girls (29, 30). Children living in families with lower socioeconomic status have a higher risk of asthma (31). Additionally, maternal smoking was another factor identified that could influence asthma development in both the mother and, to a greater degree, the child (32).

An interesting finding was that more people with asthma took a flu shot in the past 12 months than those who did not. As the Michigan BRFSS study mentioned, this could be due to children with asthma being more prone to receiving the flu shot, as their parents may be aware of the dangers that arise if a child with asthma gets the flu (7). Another interesting finding is the association of severe COVID-19 symptoms with asthma in youth. Currently, there are mixed results on the association of COVID-19 with asthma. A recent nationwide population-based cohort study with approximately 50 million people in South Korea showed that COVID-19 was associated with an increased risk of asthma onset (33). A retrospective cohort study on 27,423 children in Children’s Hospital of Philadelphia Care Network ages 1-16 from March 1, 2020, to February 28, 2021, showed that the COVID-19 diagnosis has no significant impact on asthma onset (34). However, that study did not examine whether severe symptoms of COVID-19 could impact asthma onset in children. Using national survey data, our study is the first to provide the association between severe COVID-19 symptoms and asthma risks in U.S. youth.

This study has several limitations. First, the NHIS does not contain every factor potentially associated with asthma, as its purpose is to get a general health overview of the public. This prevents us from identifying more significant variables associated with asthma development in youth. In addition, including those variables might lead to better model performance. Second, undersampling could have removed many records, which might negatively influence the model performance. Third, although the predictive models have relatively better model performance than previous studies, the models need to be further improved, for example, by including more variables. Fourth, the potential risk factors for asthma development in youth identified in this study must be further validated by longitudinal studies. Lastly, the hyperparameters for machine learning models could be better optimized in future studies, which might potentially improve the model’s performance.

Using the combined national survey data and applying different sampling techniques, this study successfully built several machine-learning models for predicting asthma development in youth, which will be very valuable for early screening and detection. The identification of additional potential risk factors (such as general health status and having received flu vaccine in the past 12 months) could aid in the early detection and prevention of asthma development among youth.

## Data Availability

All data produced are available online at https://www.cdc.gov/nchs/nhis/data-questionnaires-documentation.htm.

## Acknowledgments

The authors declare no potential conflicts of interest with respect to the research, authorship, or publication of this article. The authors received no external financial support for the research, authorship, or publication of this article. No copyrighted material, surveys, instruments, or tools were used in the research described in this article.

## Author Information

Corresponding Author: Matthew Xie, Pittsford Sutherland High School, Pittsford, NY, 14534.

Author Affiliations: ^1^Pittsford Sutherland High School, Pittsford, New York. ^2^Department of Computer Science, University of Rochester, Rochester, New York.

