## Supplemental Figures for "Predicting the Risk of Asthma Development in Youth Using Machine Learning Models"

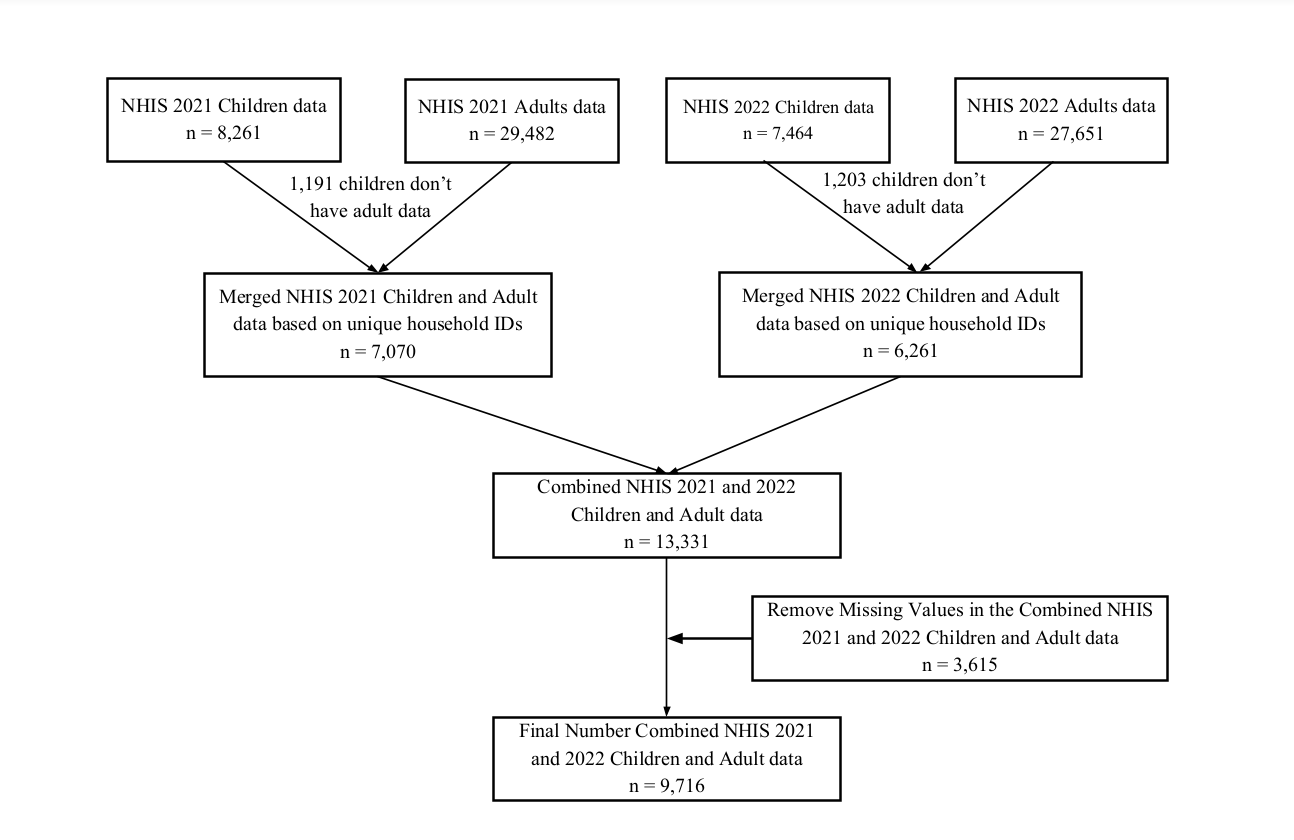

**Appendix Figure 1. NHIS combined dataset preparation**

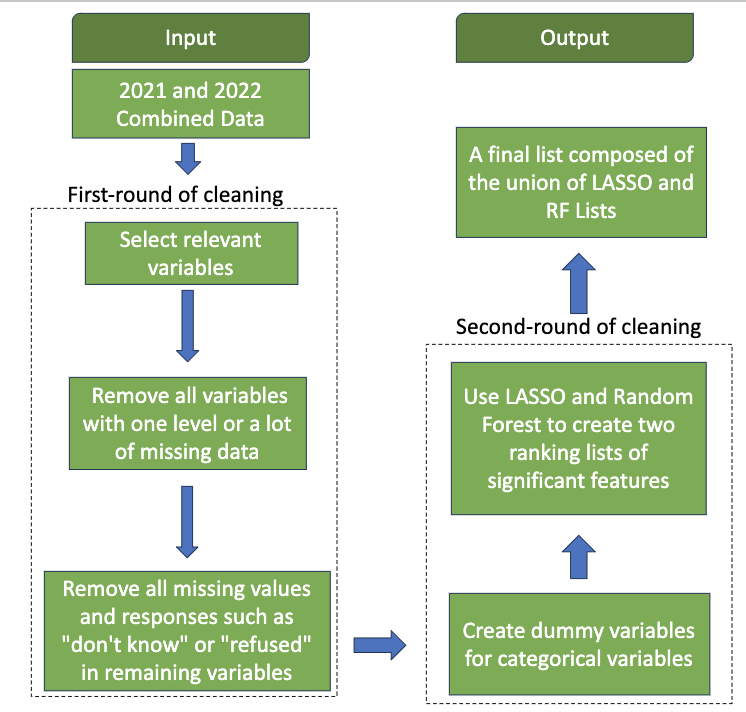

**Appendix Figure 2. Data pre-processing and feature selection**

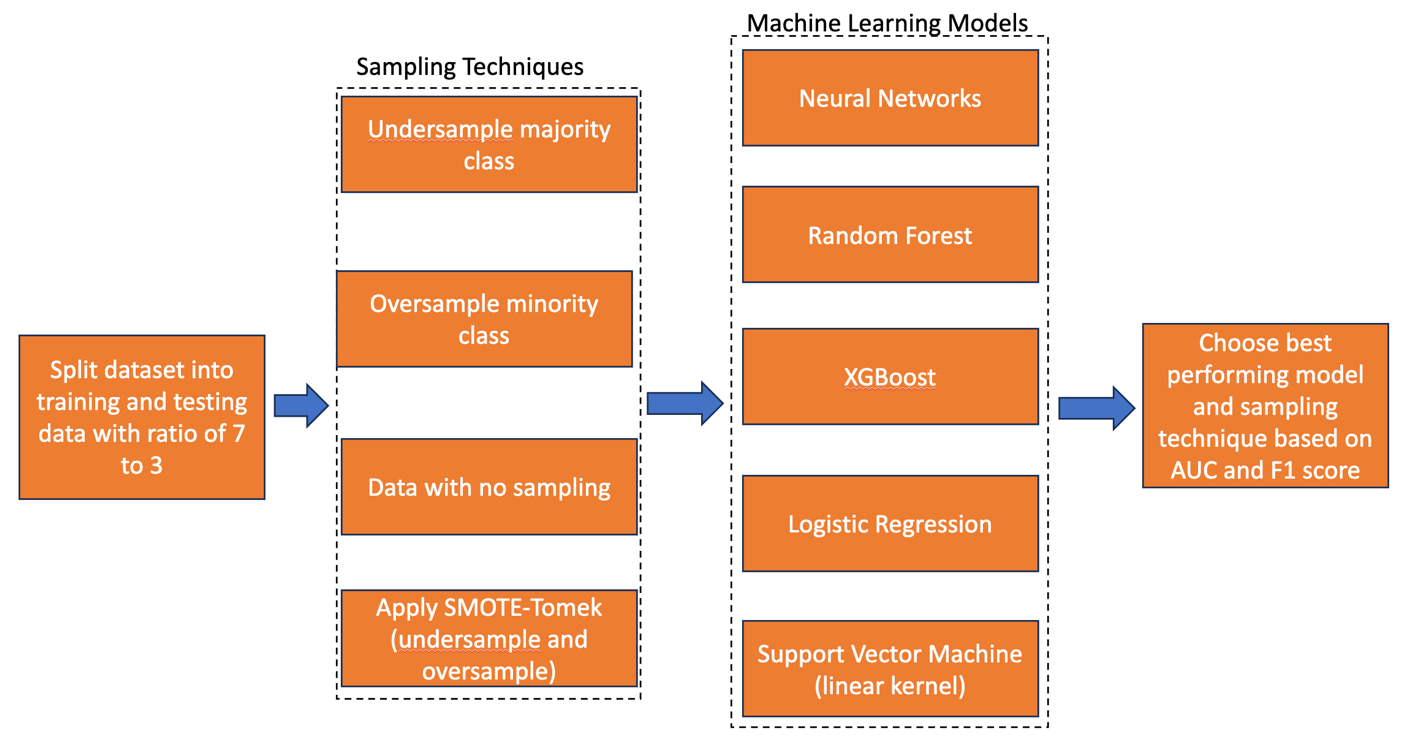

**Appendix Figure 3. Building machine learning models for asthma prediction in youth**

**Appendix Table 1. Feature selection by LASSO and Random Forest.**

| **Variables selected by LASSO** | **Variables selected by Random Forest** | **Variable Description** |
| --- | --- | --- |
| AGEP_C | AGEP_C | Age of child |
| ASEV_A | ASEV_A | Adult ever had asthma |
| EMERG12MTC_C | EMERG12MTC_C | Number of times visited emergency room in past 12 months |
| PHSTAT_C | PHSTAT_C | General Health status |
| POVRATTC_C | POVRATTC_C | Family poverty ratio |
| REGION | REGION | Household region |
| RX12M_C | RX12M_C | Took prescription medication in past 12 months |
| SEX_C | SEX_C | Gender |
| SHTFLU12M_C | SHTFLU12M_C | Received a flu shot in past 12 months |
| URBRRL | URBRRL | NCHS Urban-Rural classification |
| URGNT12MTC_C | URGNT12MTC_C | Number of times visited urgent care in past 12 months |
| ADHDEV_C |  | Ever had ADD/ADHD |
| CIGAREV_A | CIGAREV_A | Parent ever smoked a cigar |
| ALCDRUGEV_C |  | Ever lived with anyone with alcohol/drug problems |
| ASDEV_C |  | Ever had autism |
| BNEEDS_C |  | Lifetime of lacking basic needs (Yes or No) |
| COVER_C |  | Health insurance hierarchy |
| DDEV_C |  | Ever had developmental delay |
| DIBEV_C |  | Ever had diabetes |
| ECIGEV_A |  | Parent ever used electronic cigarettes |
| FDSBALANCE_C |  | How often child could not afford to eat balanced meals |
| FWIC12M_C |  | Received WIC (Women, Infants, Children) benefits in past 12 months |
| HISPALLP_C |  | Single and multiple race groups with hispanic origin |
| LDEV_C |  | Ever had learning disability |
| MEDDL12M_C |  | Delayed medical care due to cost in past 12 months |
| MENTDEPEV_C |  | Ever lived with anyone mentally ill/severely depressed |
| PIPEEV_A |  | Parent ever smoked a pipe filled with tobacco |
| PREDIB_C |  | Ever had prediabetes |
| SMKCIGST_A |  | Parent's cigarette smoking status |
| TBIDAZED_C |  | Ever dazed or memory gap |
| TBIHEADSYM_C |  | Ever had headache, vomit, blurred vision, or mood change after blow to head |
| VIOLENEV_C |  | Victim of/witnessed violence |
| WELLNESS_C |  | Was the last visit a wellness visit |
| CVDSEV_C | CVDSEV_C | Symptoms of COVID-19 |
